# A SIMULATION MODEL TO QUANTIFY THE EFFICACY OF DRY CLEANING INTERVENTIONS ON A CONTAMINATED MILK POWDER LINE

**DOI:** 10.1101/2024.08.05.24311372

**Authors:** Devin Daeschel, Long Chen, Claire Zoellner, Abigail B. Snyder

**Author notes:** Devin Daeschel and Long Chen contributed equally to this work.

## Abstract

Outbreaks of *Salmonella* and other pathogens associated with low moisture foods have been caused by cross-contamination from the processing environment into product. We used Monte Carlo simulations to model the impact of hypothetical cross-contamination scenarios of *Salmonella* from production equipment into milk powder. Model outputs include the quantity and extent of contaminated product from a production line, which can be useful for comparing the efficacy of different cleaning interventions. We also modeled the cross-contamination of potential dry cleaning surrogates to see how they responded to cleaning interventions in comparison to *Salmonella*. Input parameters for the model included log reductions from wiping an inoculated surface with a dry towel and transfer coefficients from an inoculated surface into milk powder that were measured experimentally and fitted to probability distributions. After a 2 log CFU contamination breach, the number of consumer size milk powder units (300 g) contaminated with *Salmonella* was 72 [24, 96] (median [p5, p95] across 1000 simulation iterations). The average concentration of *Salmonella* within contaminated units was -2.33 log CFU/g [-2.46, -1.86]. Wiping with a dry towel reduced the number of contaminated units to 26 [12, 64]. After product flushing with 150 kg of milk powder, the number of contaminated units dropped to 0 [0, 41]. *E. faecium* was the most appropriate surrogate for *Salmonella* transfer from surface to milk powder, while *L. innocua* was a more appropriate surrogate for the dry towel wiping intervention. These results suggest that product flushing, and to a lesser degree dry wiping, may be effective interventions in reducing contaminated milk powder product after a contamination breach. Further, simulation modeling is a useful tool for evaluating *Salmonella* dry transfer surrogates for their use in dry cleaning validation and modeling applications.

**IMPORTANCE:** This work demonstrates the utility of *in silico* modeling as a decision support tool that can 1) estimate the cross-contamination of *Salmonella* into milk powder under different processing scenarios, 2) compare the efficacy of different cleaning interventions and 3) help inform surrogate selection for the dry transfer of *Salmonella* in modeling and cleaning validation applications. The model presented here contributes to the risk-benefit analysis of tradeoffs associated with dry cleaning in low moisture food environments. For example, the model can be applied to estimate the efficacy of cleaning interventions like product flushing at a lower resource cost than experimental trials in a processing line. The model presented here also provides a more interpretable metric for choosing appropriate *Salmonella* surrogates for dry cleaning validation.

## 1. INTRODUCTION

Recent outbreaks in powdered infant formula, cereals, and peanut butter [1] as well as numerous outbreaks of *Salmonella* in milk powder [2] have underscored the challenge of environmental cleaning and sanitation in low moisture food facilities. *Salmonella* is of particular concern since it can persist in low moisture processing environments [3], cross-contaminate low moisture foods [4], and then persist in contaminated food products [5]. Epidemiological investigations of past powdered milk outbreaks are frequently inconclusive, but some have identified processing line equipment to be a likely source of contamination [3, 6]. The U.S. Food and Drug Administration (FDA) conducted a sampling survey of 55 milk powder processing facilities and detected *Salmonella* species in 6% of facilities [7]. Despite best efforts to maintain a completely dry processing line, water breaches (e.g. faulty plumbing, roof leaks, floods, leftover water from wet cleanings) are common and may introduce contamination or promote the growth of pathogens already in the environment.

Milk powder processors rely on dry cleaning strategies such as brushing, vacuuming, and product flushing to maintain a hygienic processing environment without introducing water.

Flushing (also referred to as product push-through, dry purging, or dry rinsing) is a sanitation strategy in which product or another dry material is run through the processing line and discarded to remove food, allergenic residues, or in some cases microbial contaminants [8, 9]. These practices are effective at removing visible food residues and are an integral part of the environmental sanitation process [10]; however, research on the efficacy of these techniques for microbial removal and reducing the risk of cross-contamination is currently limited [11]. As a result, FDA does not consider dry cleaning methods alone to be a valid sanitation break in low moisture food (LMF) processing [12]. On the other hand, the efficacy of aqueous sanitizers for microbial reduction is well established [13–15], but the increased risk of microbial growth from the introduction of water to a LMF processing environment remains a significant trade-off.

Therefore, there is currently a knowledge gap around the efficacy of dry cleaning at reducing the risk of microbial cross-contamination and the risk trade-offs between dry and wet sanitation processes used in milk powder processing. Computer modeling as a tool to estimate risk trade-offs as been applied to other food products [16–22], but application to milk powder is currently limited [23]. For milk powder, digital tools that can model the cross-contamination of pathogens after a contamination breach can be broadly helpful for various applications. For example, to estimate the efficacy of different cleaning and sanitation strategies used in industry, or to estimate the number of contaminated servings a population may be exposed to as part of a Quantitative Microbial Risk Analysis (QMRA) [24].

The goals of this study were to 1) build a simulation model of environmental *Salmonella* transfer into milk powder during a production run, 2) estimate the prevalence and concentration of *Salmonella* in contaminated milk powder units produced during a production run after a contamination breach, 3) estimate the effects of product flushing and dry towel wiping on the prevalence and concentration of contaminated milk powder units, 4) estimate the amount of cross-contamination from a cleaning tool used on a contaminated surface and 5) evaluate potential dry cleaning surrogates for *Salmonella*.

## 2. MATERIALS AND METHODS

### 2.1 Modeling a milk powder production run

Milk powder production in a dry food processing environment was modeled in this study. Processing parameters for the production run were chosen to reflect those of a typical, industrial milk powder manufacturer. Each modeled production run produced 30,000 kg of milk powder, partitioned into 100,000 units, each containing 300 g of milk powder (consumer-sized units). Milk powder production typically involves pasteurization of liquid milk, concentration by evaporation or reverse osmosis, formation of powder via spray drying, and then transportation and packaging of the final product [25]. The transportation step between powder formation and packaging (i.e. dry powder flowing through a pipe or over a surface) was chosen as the site of the contamination breach and source of subsequent cross-contamination. The main outputs from the model are the number of milk powder units that were contaminated during the production run (prevalence), and the mean concentration of contamination (log CFU/g) within the contaminated units. All modeling and statistical analysis was done in R version 4.2.2 [26]. Each model was simulated with 1,000 iterations, with each iteration representing a single production run. Results for each simulated scenario are reported as the median [5^th^, 95^th^ percentile] of the models outputs of 1,000 iterations. Preliminary testing showed that increasing the number of iterations did not meaningfully change the model outputs.

Five different *Salmonella* cross-contamination and intervention scenarios were modeled (Fig. 1). In scenario 1, a stainless-steel food contact surface on the production line, such as the filler pipe or transportation equipment, is contaminated with *Salmonella* with no intervention prior to a production run. Three hypothetical initial surface contamination levels (2 log CFU, 4 log CFU, 6 log CFU) were tested. Scenarios 2 and 3 incorporated different dry cleaning strategies prior to the start of production (product flushing and wiping with a dry towel, respectively). Scenario 4 models introduction of contamination from the cleaning intervention tool, a dry towel. In this scenario, a dry paper towel is used to clean a contaminated surface (6 log CFU), and then used again on a surface of the production line. Scenario 5 simulated a more rigorous dry cleaning intervention, repeated wiping of the contaminated surface with a dry towel, to estimate how many passes of the towel are needed to remove all the surface contamination prior to production. These estimates were compared across potential bacterial surrogates (*E. coli*, *E. faecium*, *L. innocua*) for *Salmonella* and dry cleaning validation and verification activities.

**Figure 1:**
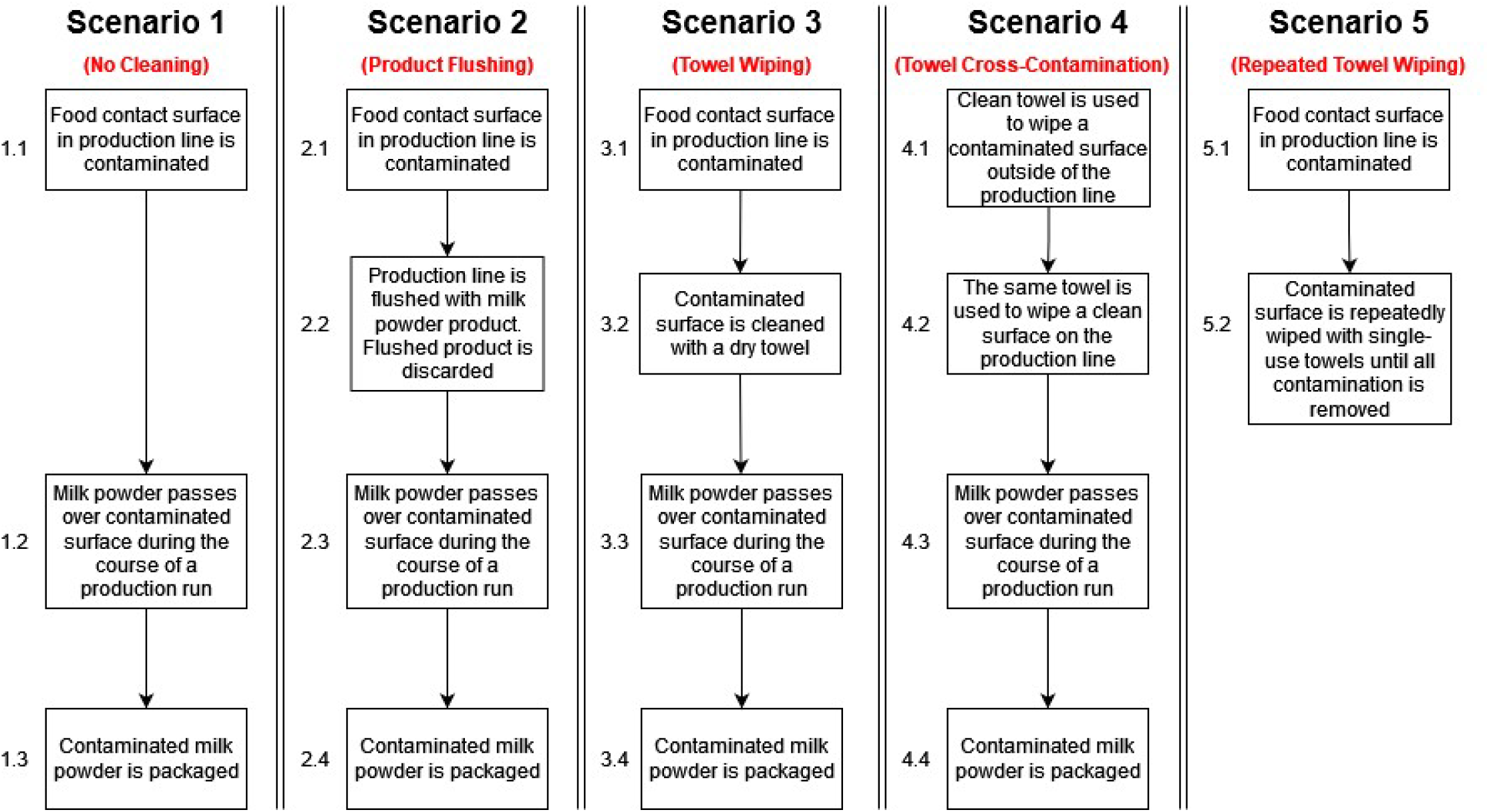
The different scenarios considered in the model. Scenario 1 (cross-contamination from a contaminated surface into milk powder), scenario 2 (cross-contamination from a contaminated surface into milk powder after a product flushing intervention), scenario 3 (cross-contamination from a contaminated surface into milk powder after a dry towel wiping intervention), scenario 4 (cross-contamination from a contaminated towel, to a stainless-steel surface, and then into milk powder), and scenario 5 (repeated towel wiping of a contaminated stainless-steel surface without production of milk powder).

### 2.2 Modeling initial contamination

Each iteration in the base model starts with a user-defined quantity of initial contamination on the processing line. There is limited published research enumerating *Salmonella* from food processing surfaces in real-world or realistic scenarios, but one study reported that the *Salmonella* contamination on processing surfaces averaged 1.3 CFU/cm^2^ after a production run with contaminated dry poultry feed [27]. Target aerobic plate count values for food processing surfaces may be set as hygienic indicators. These values typically vary depending on the environment being sampled, but less than 500 CFU prior to sanitization is recommended as acceptable by the Almond Board of California [28]. We chose 2 log, 4 log, and 6 log CFU as starting contamination loads introduced on the filler pipe surface to encompass a range of hypothetical initial contamination levels. The 2 log CFU load was considered the most likely initial contamination level and was used as the primary point of comparison between scenarios.

### 2.3 Modeling microbial transfer and reduction

The main function of the simulation models the transfer of bacteria from a contaminated stainless-steel surface into milk powder. Transfer data for *S. enterica, E. faecium, L. innocua*, and *E. coli* from a spot inoculated stainless-steel coupon into 10 g quantities of milk powder were generated in the present work (described below, data available in supplementary file 1).

Probability distributions (Table 1) were fit to the transfer data using the fitdist function in the ‘fitdistrplus’ package version 1.1.8 [29]. For transfer from a spot inoculated surface to milk powder, transfer coefficients were log-transformed and then fitted to a normal distribution [30, 31]. Goodness of fit was measured with the Kolmogorov–Smirnov test and Q-Q probability plots. Simulated production runs were modeled through the following steps for each iteration:

1) A user defined quantity of starting contamination (*C_0_*) on the stainless-steel filler pipe is selected:

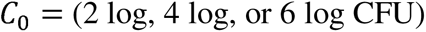

2) A random log-transformed transfer coefficient (*T_c_*) is sampled from the surface to milk powder log-normal transfer coefficient distribution:

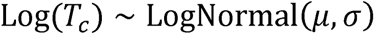

3) The sampled transfer coefficient is used in a binomial distribution to calculate the number of CFU transferred (*N_t_*) into each 10 g quantity of milk powder that contacts the contaminated surface at contact event *t*, where *t* is each contact event between a 10 g quantity of milk powder and the contaminated surface.

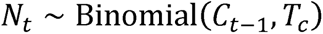

4) The remaining contamination on the surface (*C_t_*) is updated to reflect the CFU that was transferred in the previous contact event:

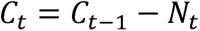

5) Steps 3 and 4 are repeated until all milk powder (30,000 kg) is run through the production line or until all contamination on the surface has been transferred (*C_t_* = 0).

**Table 1:**
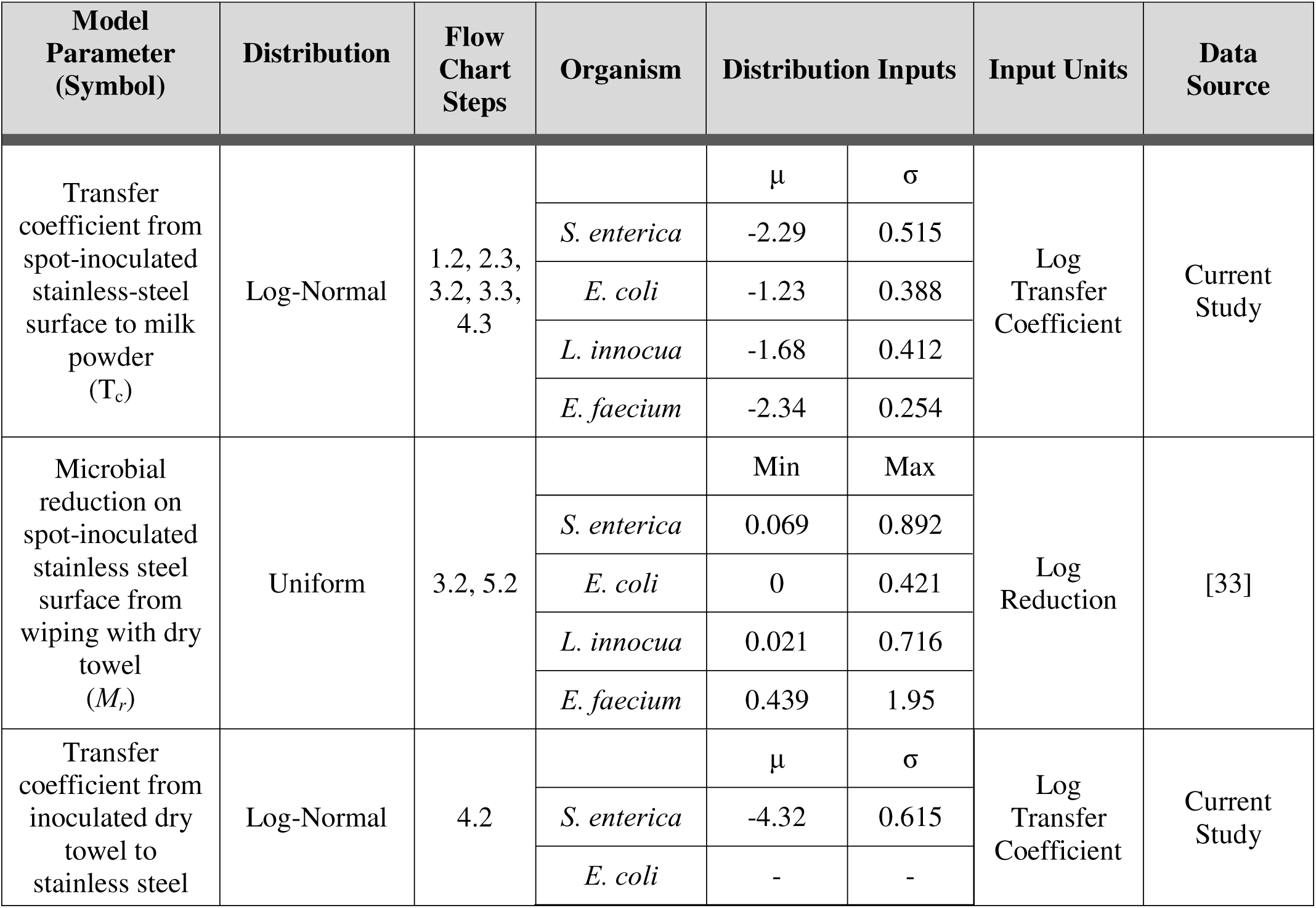
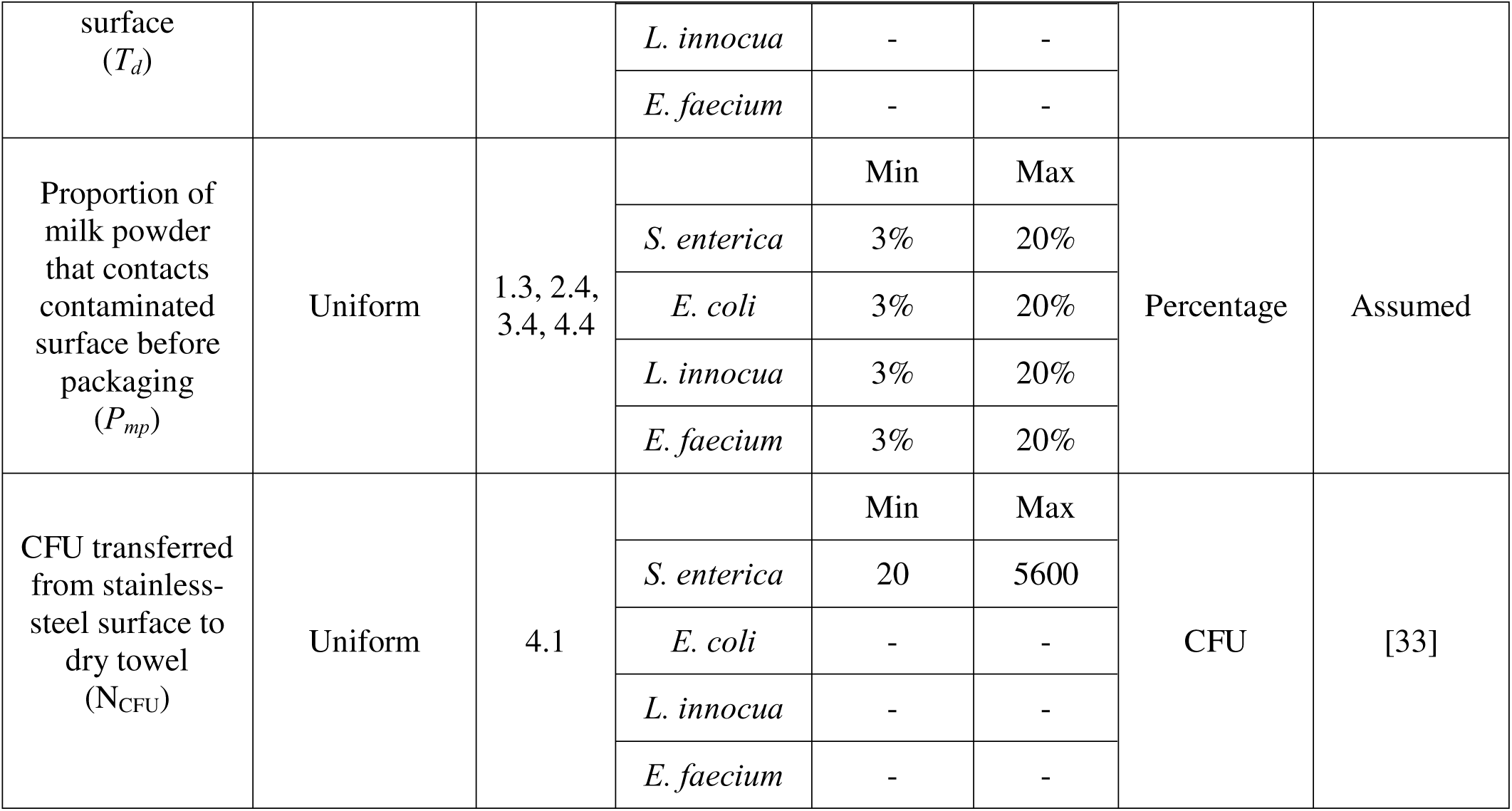
Model input parameters, distributions, and references for microbial transfer and reduction during milk powder processing.

It is unlikely that the entire quantity of milk powder moving through a processing line would contact a single contaminated surface site. Only a fraction of milk powder moving through the line would have the chance to contact the contaminated surface and for microbial transfer from the surface to occur. This same principle would apply to a niche or “dead zone” on the processing line which the milk powder product only infrequently contacts. Therefore, we implemented a parameter in the model to account for the proportion of milk powder in a single product unit (300 g) that contacts the contaminated surface before being packaged. We assumed that anywhere from 3% (10 g) to 20% (60 g) of the milk powder entering a product unit would contact the contaminated surface. Since microbial transfer was calculated for each 10 g of milk powder passing through the line, a single milk powder product unit (300 g) could contain a minimum of 10 g and a maximum of 60 g of contaminated milk powder, representing 1 to 6 total transfer events for all milk powder in that unit, respectively. At the end of an iteration, the proportion of milk powder (*P_mp_*) in each unit that contacted the contaminated surface is sampled from a uniform distribution:

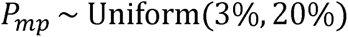

The sampled proportion was then used to calculate the number of CFU in each milk powder unit. This stochasticity was modeled with a uniform probability distribution, which is typical in situations where there is no *a priori* data [32].

A fourth parameter was added to model scenario 3 (wiping with a dry towel intervention). This parameter represented the microbial reduction (*M_r_*) when a dry towel is used to wipe the contaminated surface. This was described using a uniform distribution with minimum and maximum values taken from microbial reduction data (Table 1)

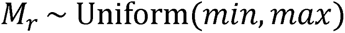

Microbial reduction data for *Salmonella* on a spot inoculated stainless-steel coupon wiped with a dry towel, and data on the microbial transfer to the towel, was previously reported by Chen et al. [33] (Supplementary file 1).

For scenario 4, an additional parameter (*T_d_*) for the transfer from a contaminated dry towel to a clean surface was included using a transfer coefficient log-normal probability distribution (Table 1):

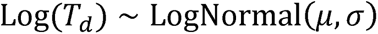

Transfer data for *S. enterica* on an inoculated dry towel to a clean stainless-steel surface was generated in the current study (Supplementary file 1). In scenario 4, the dry towel initially became contaminated from being used to wipe a contaminated surface outside of the modeled processing line. The amount of contamination on this initial surface was set to 6 log CFU to represent a worst-case scenario. The amount that gets transferred to the dry towel from the surface was modeled using a uniform distribution:

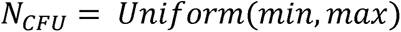

Minimum and maximum amounts of CFU transferred from the surface to the towel were reported by Chen et al. [33].

### 2.4 Sensitivity analysis

Starting contamination (*C_0_*), transfer coefficient from stainless-steel surface to milk powder (*T_c_*), microbial reduction during dry cleaning (*M_r_*), and the proportion of milk powder that contacted the contaminated surface within each unit (*P_mp_*) were included as parameters in a global sensitivity analysis to determine their relative effects on the prevalence and concentration of contaminated milk powder units during a scenario 3 (towel wiping) production run. Scenario 3 was chosen for the sensitivity analysis so that microbial reduction from wiping with a dry towel could be included as a parameter. Sensitivity analysis was performed using partial rank correlation coefficients (PRCC) and was conducted with the ‘epiR’ package version 2.0.66 in R [34]. The model makes several necessary simplifications that may affect how representative the model is compared to real-world scenarios. For example, we assume that the rate of transfer is independent of the starting contamination and that bacterial transfer events are independent of each other, which is typical of similar models of microbial transfer [30, 31]. We also did not implement the possibility of contamination being spread or re-introduced in the processing line from cross-contaminated milk powder. In other words, *Salmonella* could not transfer to a new surface from the milk powder itself. For a given production run we also assumed that the sampled transfer coefficient and proportion of milk powder in each unit that contacted the contamination were constant for the duration of that simulation. Finally, the intervention using dry towels was done with three passes of the towel, each with a standardized sheer-stress which, while experimentally consistent, may not be accurate to real life wiping done by a human.

### 2.5 Bacterial strains

The same bacterial strains were used in our previous study associated with dry sanitation [33]. The *Salmonella* strain chosen for this study was *Salmonella* Enteritidis PT30 since it has been associated with an outbreak and has previously been used in low moisture food research.

The surrogate strains used in this study were *Enterococcus faecium* NRRL B-2354, *Listeria innocua* ATCC 51742 and *Escherichia coli* ATCC 25922. The *E. faecium* strain was chosen because it has been used as a thermal processing surrogate for *Salmonella*. The *L. innocua* and *E. coli* strains were chosen since both have been used in prior surface attachment studies.

### 2.6 Inoculum preparation

The same inoculum preparation method was used as described in Chen and Snyder (2023). Briefly, a loopful of frozen stock for each bacterium was inoculated into Brain Heart Infusion (BHI) broth (BD, Thermo Fisher Scientific, Waltham, MA, USA) and incubated at 37°C for 24 h. Overnight broth suspensions were streaked onto BHI agar plates and incubated at 37°C for 24 h. An isolated colony was transferred from stock plates into BHI broth followed by incubation at 37°C for 24 h. After incubation, the culture was centrifuged (Eppendorf 5804R, Eppendorf, NY, USA) at 10,000 RPM for 5 min, and the cell pellet was washed twice in 0.1% phosphate buffered saline (PBS) (BD, Thermo Fisher Scientific, Waltham, MA, USA). After washing, the cell pellets were resuspended in the same volume of 0.1% PBS, to achieve a cell concentration ∼9.0 log CFU/mL. Inoculation procedures were repeated for each of the four bacteria using three biological replicates.

### 2.7 Surface inoculation and microbial transfer

Microbial reduction of an inoculated stainless-steel surface wiped with a dry towel, and transfer from the inoculated surface to the towel were measured previously by Chen et al. in triplicate [33]. Transfer coefficients from an inoculated stainless-steel surface to milk powder and for an inoculated dry towel to a stainless-steel surface were generated as follows:

#### i) Stainless-steel surface to milk powder

Eighteen 10 μL drops of inoculum were spot inoculated on sanitized stainless-steel coupons (2.4 cm in width, 3.5 cm in length, and 0.48 cm in thickness, AISI 304 stainless-steel 2B finish). Then, the coupons were dried in a biosafety cabinet overnight with the fan on (Thermo Fisher Scientific, Waltham, MA, USA). Each inoculated coupon was manually shaken and mixed with 10 g of milk powder (Nestle Carnation, Switzerland) in a sterile sample bag for 5 min to facilitate the microbial transfer from the inoculated coupon to milk powder. Cells were collected and enumerated from: (1) coupons before the transfer treatment to milk powder, (2) coupons following the transfer treatment, and (3) the milk powder following the transfer treatment.

Briefly, coupons were rinsed with 10 mL 0.1% PBS in sample bags (Whirl-Pak™, Madison, WI, USA), and 10 g milk powder was diluted with 20 mL 0.1% PBS in sample bags. Samples were diluted and spread plated on Brain Heart Infusion (BHI) (BD, Thermo Fisher Scientific, Waltham, MA, USA) and then enumerated. Microbial transfer experiments were conducted for 5 replicates for each bacterium.

#### ii) Dry towel to stainless-steel surface

A dry towel was inoculated as follows: Glass beads were first inoculated with each of four bacterial strains according to the same method described by Chen et al. [33]. Autoclaved dry paper towels were inoculated through manual mixing and rolling with glass beads (Walter Stern Inc., Port Washington, NY, USA) for 1 min. Stainless-steel coupons were first sanitized in 10 % bleach for 5 min, then wiped with dry towels, soaked in 70% ethanol for 2 min, wiped with a dry towel, then air dried overnight before treatment. Microbial transfer from an inoculated dry towel to stainless-steel coupons was done through a simulated dry cleaning process described by Chen et al. [33]. Cells were collected from untreated dry towels, coupons following the cleaning treatment, and the dry towel following the cleaning treatment and enumerated with serial dilution and spread plating on Brain Heart Infusion (BHI) (BD, Thermo Fisher Scientific, Waltham, MA, USA). Briefly, dry towels were diluted with 10 mL 0.1% PBS in sample bags (Whirl-Pak™, Madison, WI, USA) and coupons were diluted with 2 mL 0.1% phosphate buffered saline (PBS) (BD, Thermo Fisher Scientific, Waltham, MA, USA) in sample bags which yielded a detection limit of 0.30 log CFU. Microbial transfer experiments were conducted for 5 replicates for each bacterium.

## 3. RESULTS AND DISCUSSION

### 3.1 Transfer coefficient and the proportion of milk powder that contacted the contaminated surface were highly correlated with product contamination outcomes

A global sensitivity analysis for scenario 3 was conducted to assess the relative importance of model parameters on the prevalence and concentration of *Salmonella* in contaminated milk powder units (Fig. 2). The parameters most correlated with both prevalence and concentration of *Salmonella* were the starting contamination (*C_0_*), the transfer coefficient from stainless-steel to milk powder (*T_c_*), and the proportion (*P_mp_*) of milk powder in each product unit that contacted the contaminated surface. Both the transfer coefficient and the proportion of milk powder contacting the contamination were positively correlated with concentration, but negatively correlated with prevalence. This is because a higher transfer coefficient and a higher proportion of milk powder contacting the contaminated surface results in *Salmonella* transferring at greater level into a smaller number of product units. Indeed, the transfer coefficient had almost as much impact on outcomes as the amount of starting contamination (2 log, 4 log, or 6 log CFU). And the proportion of milk powder in a unit that contacted the contaminated surface was more impactful than whether or not there was a dry wiping intervention (Fig. 2).

**Figure 2:**
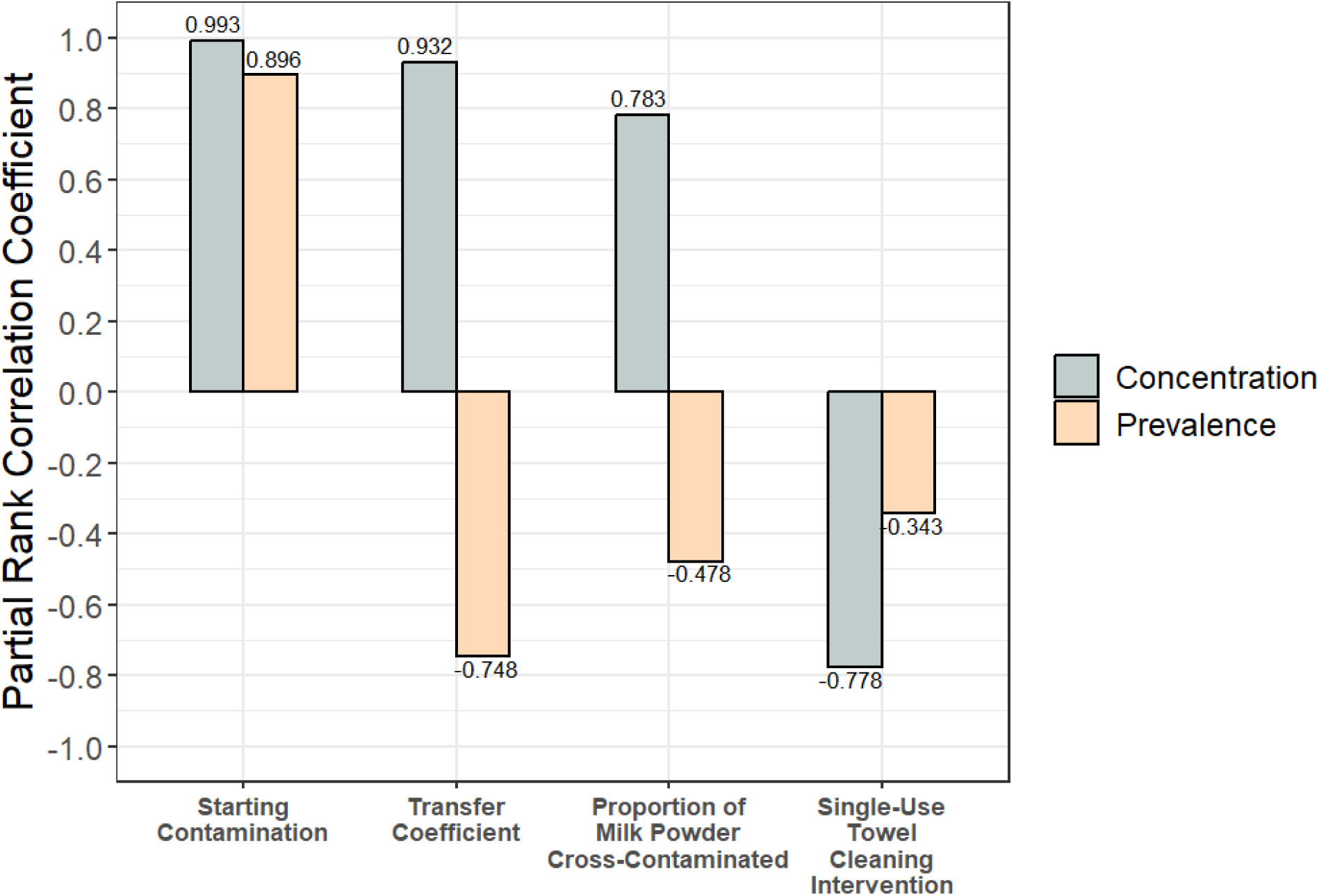
Partial rank correlation coefficients for all model input parameters were correlated with the prevalence and concentration of contaminated units (scenario 3). *The sampled transfer coefficient had a large effect on concentration and prevalence*.

The transfer coefficient being highly correlated with the prevalence and concentration of contaminated units underscores the importance of accurate and representative data used for this parameter. Previous works have shown that various factors can affect microbial transfer: inoculation method [33], contact time between the contaminated surface and food product [35], surface material [36, 37], and pathogen strain selection [38]. Future research on designing experimental transfer conditions to be as realistic to processing conditions as possible will be essential to generate accurate transfer rates for modeling applications and to inform surrogate selection. Similarly, the proportion of milk powder that contacted the contaminated surface was highly influential on model outputs. Our current model represents only one simplified example of how a contamination breach could manifest (on a spot-inoculated stainless-steel surface that milk powder passes over on its way to packaging). However, in real-world line breaches, contamination may be harbored in parts of the line that contact milk powder more or less frequently than what we considered in our model. For example, the interior of a spray drier possesses many niches that are difficult to clean and may have unique cross-contamination dynamics. One study found *Salmonella* contamination in all tested areas of a spray drier after inoculated soy protein isolate was ran through it [39]. Additionally, different niches on the interior of a spray drier may experience different levels of shear stress from product moving through the line which could impact the rate and frequency of microbial transfer. This may be further complicated by equipment vibration or aerosolization, and contamination distributed across more surface area could result in more milk powder contacting the contaminated surface. Therefore, a complete understanding of the cross-contamination dynamics in different parts of the processing line will be critical for generating accurate model parameters. Collaboration with industry stakeholders to better understand how surface contamination interacts with product in problematic niches or “dead zones” within active processing lines will help towards this end.

### 3.2 A contamination breach in a milk powder processing line resulted in a low number of contaminated units and a low concentration of *Salmonella* within those units

Scenario 1 (Fig. 1) was simulated 1,000 times to quantify the prevalence and concentration of milk powder units contaminated with *Salmonella* during a hypothetical processing line breach where 2 log CFU of *Salmonella* was introduced onto a surface. The number of contaminated units across all simulations was 72 [24, 96] (Fig. 3A). The average concentration of *Salmonella* within contaminated units was -2.33 log CFU/g [-2.46, -1.86] (Fig. 3B). When the contamination breach was increased to 6 log CFU *Salmonella*, the number of contaminated units increased to 688 [95, 4420], a 2.9 log percent increase compared to the 2 log CFU initial contamination level (Fig. 3C). Similarly, the average concentration of *Salmonella* within contaminated units for a simulated production run increased to 0.689 log CFU/g [-0.122, 1.55] (5 log percent increase) (Fig. 3D). While both the number of contaminated units and the average concentration of *Salmonella* within contaminated units increased with higher initial contamination levels, the concentration increased more than the prevalence. This was also apparent in the sensitivity analysis that showed a stronger correlation between starting contamination level and concentration than between starting contamination level and prevalence (Fig. 5). Notably, the variability in the number of contaminated products in each simulated production run also increased between the 2 log (Standard deviation: 23 product units) and 6 log (Standard deviation: 2523 product units) contamination scenarios. Similarly, the standard deviation in average concentration also increased from 0.20 to 0.52 log CFU/g. This is because as the amount of contamination increases, so does the possible number of contaminated units and the concentration of contamination within those units.

**Figure 3:**
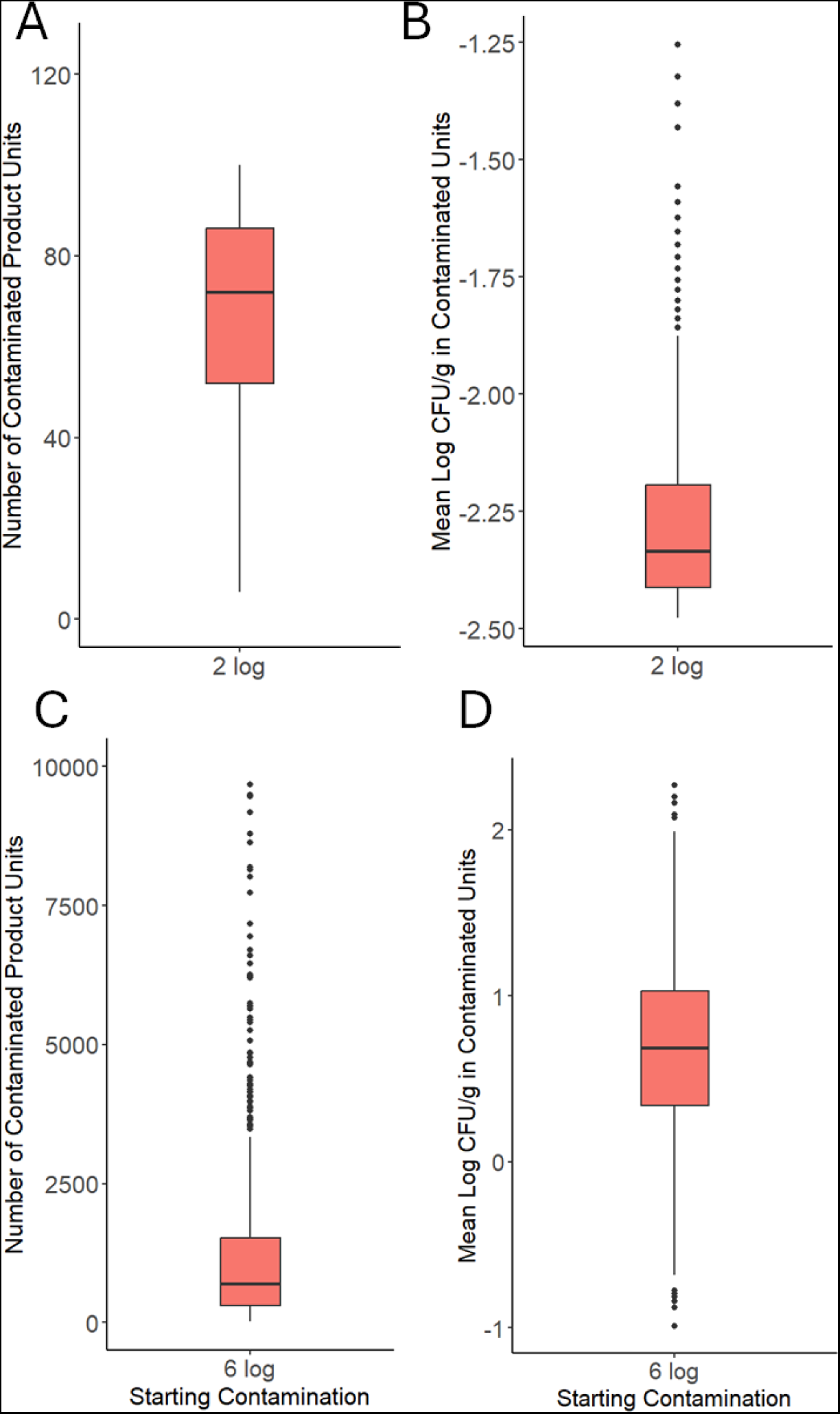
The results from 1,000 simulated production runs (scenario 1: no cleaning) are plotted for 2 log CFU **(A and B)** and 6 log CFU **(C and D)** starting contamination levels. **(A and C)** Each data point represents the number of milk powder units (300 g) contaminated with *Salmonella* in a simulated production run. **(B and D)** Each data point represents the average concentration of *Salmonella* within the contaminated milk powder units of a simulated production run.

Data on the exact number of contaminated product units during outbreaks or recalls is limited; however, there are studies that have enumerated the concentration of pathogens in recalled product units (Table 2). These studies have estimated the amount of contamination within recalled dry powder products to be between -2.82 and -1.62 log CFU/g (Table 2). This concentration range was similar to what was predicted by our model during a 2 log CFU *Salmonella* breach: -2.33 log CFU/g [-2.46, -1.86] (Fig. 3B). Notably, these studies included *Cronobacter* spp., *Salmonella* Ealing, *E. coli* O121, and *Salmonella* Typhimurium in powdered infant formula, wheat flour, and baking mix [40–43]. More experimental data is necessary to validate this model, especially with regard to the predicted number of contaminated units produced after a contamination breach.

**Table 2:** Summary of past studies that have enumerated the concentration of pathogens in recalled dry powder products. Concentrations reported by studies were converted to log CFU/g or log MPN/g for ease of comparison. *The reported concentrations of pathogens in recalled products were similar to what our model predicted during a contamination breach*.

| Recalled Product Details | Organism | Reported Contamination Concentration | Converted Concentration (log CFU/g or log MPN/g) | Reference |
| --- | --- | --- | --- | --- |
| Powdered infant formula produced in January 2007. A 22,000 kg batch was recalled. Product was packaged and sold as two 400 g bags. | *Cronobacter* spp. | -2.78 log CFU/g | -2.78 log CFU/g | [40] |
| Powdered infant formula produced in 1985. Product was packaged and sold in 25 kg bags. | *Salmonella* Ealing | 1-6 CFU/450 g | -2.65 to -1.87 log CFU/g | [41] |
| Wheat flour produced in 2016. Product was packaged and sold in 1kg and 10kg bags. | *E. coli* O121 | 0.15 to 0.43 MPN/100 g | -2.82 to -2.37 log MPN/g | [42] |
| Raw wheat flour used in baking mix produced in 2008. | *Salmonella* Typhimurium | 0.0036 to 0.024 MPN/g | -2.44 to -1.62 log MPN/g | [43] |

The predicted prevalence and concentration of contaminated milk powder units can be used to contextualize food safety risks. Assuming a 25 g serving size for milk powder, the single most contaminated product unit across all 2 log CFU simulations was only 5.5 CFU/25 g. As a point of comparison, conservative models of *Salmonella* infectious dose have predicted an infection ID50 as low as 7 CFU [44]. Notably, any amount of ingested *Salmonella* has the potential to cause illness, and heterogenous distributions of bacteria within contaminated product units could result in some servings having a higher pathogen concentration. One study enumerating *Cronobacter* from recalled powdered infant formula found that the average contamination concentration in recalled formula was -2.78 log CFU/g, but clusters as high as 2.75 log CFU/g were detected [40]. Overall, this suggests that the average contamination concentration of *Salmonella* in recalled dry powder products like milk powder is likely low for the majority of contamination breaches (< 2 log CFU). Contamination breaches may therefore go unnoticed as they result in concentrations that fall below the limit of detection in 25 g product samples [45], are present in a fraction of all units produced within a lot, and are below the level likely to cause illness. However, our model also showed that if the quantity of *Salmonella* in a breach was increased to 6 log CFU, then the *Salmonella* concentration within contaminated units increased to levels with more potential to cause illness. Breaches of this level are probably less likely to occur but could potentially arise from a severe scenario like a flood or roof leak where contamination and moisture are introduced.

### 3.3 Product flushing reduced the prevalence and concentration of contaminated units after a contamination breach

Milk powder units produced directly after the contamination breach (scenario 1) had a higher concentration of *Salmonella* compared to units produced later in the production run (Fig. 4). The concentration of *Salmonella* in the first unit produced after a 2 log CFU contamination breach was 0.01 CFU/g [0, 0.04], whereas the 100^th^ unit had a contamination concentration of 0 CFU/g [0, 0.003] (Fig. 4). For a 6 log CFU contamination breach, the first and 100^th^ product units had contamination concentrations of 108 CFU/g [6.93, 424] and 6.97 CFU/g [0, 12.3], respectively (Fig. 4). This was a result of the initial contamination rapidly transferring from the surface into the initial product units, which left fewer cells on the surface to transfer into units later in production. The industry often utilizes this phenomenon by applying dry flushing to processing lines.

**Figure 4:**
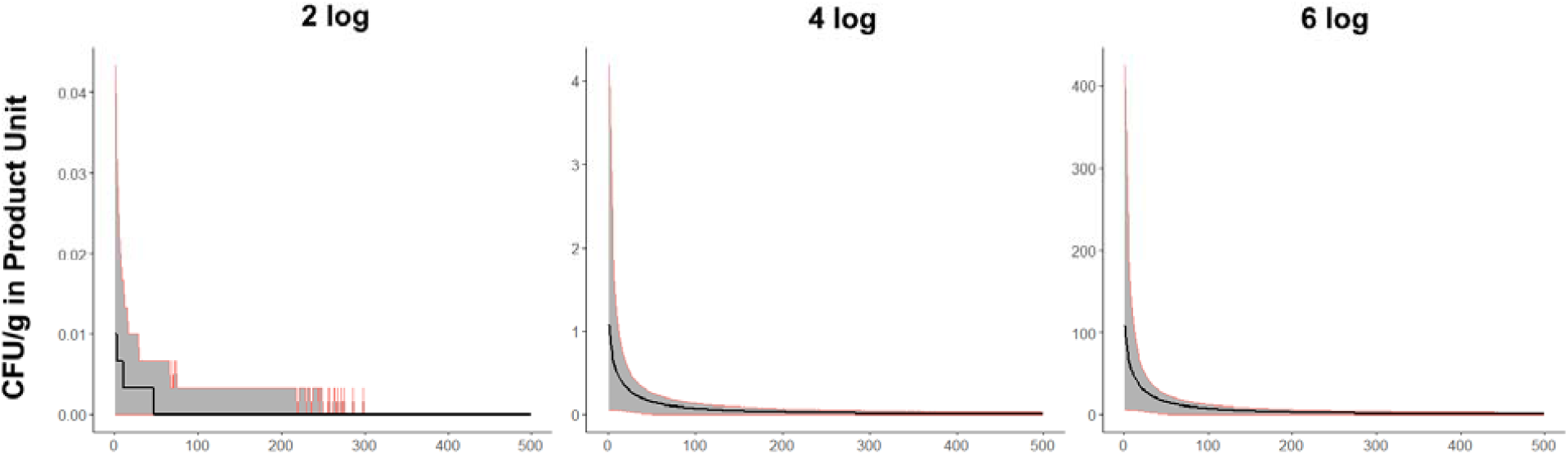
Across 1,000 simulated production runs (scenario 1), the median contamination concentration (CFU/g) in the n-th milk powder unit (i.e. from the 1^st^ unit produced in the production run or the 500^th^) is graphed in black with 5^th^ and 95^th^ percentiles graphed in red. *The concentration of contamination within product units decreased exponentially as sequential units were produced in a simulated production run*.

To evaluate the efficacy of this intervention, we simulated product flushing (scenario 2) with variable amounts of milk powder (30 kg, 150 kg, and 300 kg). Increasing the amount of flushed material resulted in fewer finished products with *Salmonella* (Fig. 5). Without any product flushing (scenario 1) the median number of contaminated units across all simulations was 72 [24, 96]. This was reduced to 20 [0, 82] contaminated units after flushing with 30 kg, and further reduced to 0 [0, 41] units after flushing with 150 kg. Flushing with 300 kg resulted in 0 [0, 16] contaminated units. Flushing with 30 kg, 150 kg, and 300 kg resulted in 17.4%, 63% and 79.9% of simulated production runs having no contaminated units, respectively. These results suggest that product flushing directly after a suspected breach may quickly remove the majority of contaminating cells and reduce the amount of downstream contaminated product. However, the variability in outputs between flushing simulations should also be considered. Even the most rigorous flushing intervention (300 kg flush) after a 2 log breach had a minority of simulations resulting in more than 50 contaminated product units (Fig. 5). Additionally, current limitations in the existing model should be further explored. For example, “dead spots” in the processing line that do not come into consistent contact with the flushing material must be controlled. These dead spots may act as a niche or reservoir for contamination. The sensitivity analysis showed that the amount of milk powder contacting the contaminated surface is highly correlated with product contamination outcomes. Accordingly, this parameter also influences how much *Salmonella* contamination is removed during product flushing. If the contaminated surface is located in an area of the processing line that frequently contacts milk powder, then flushing will be more effective at removing it. Additionally, flushing has economic and sustainability tradeoffs.

**Figure 5:**
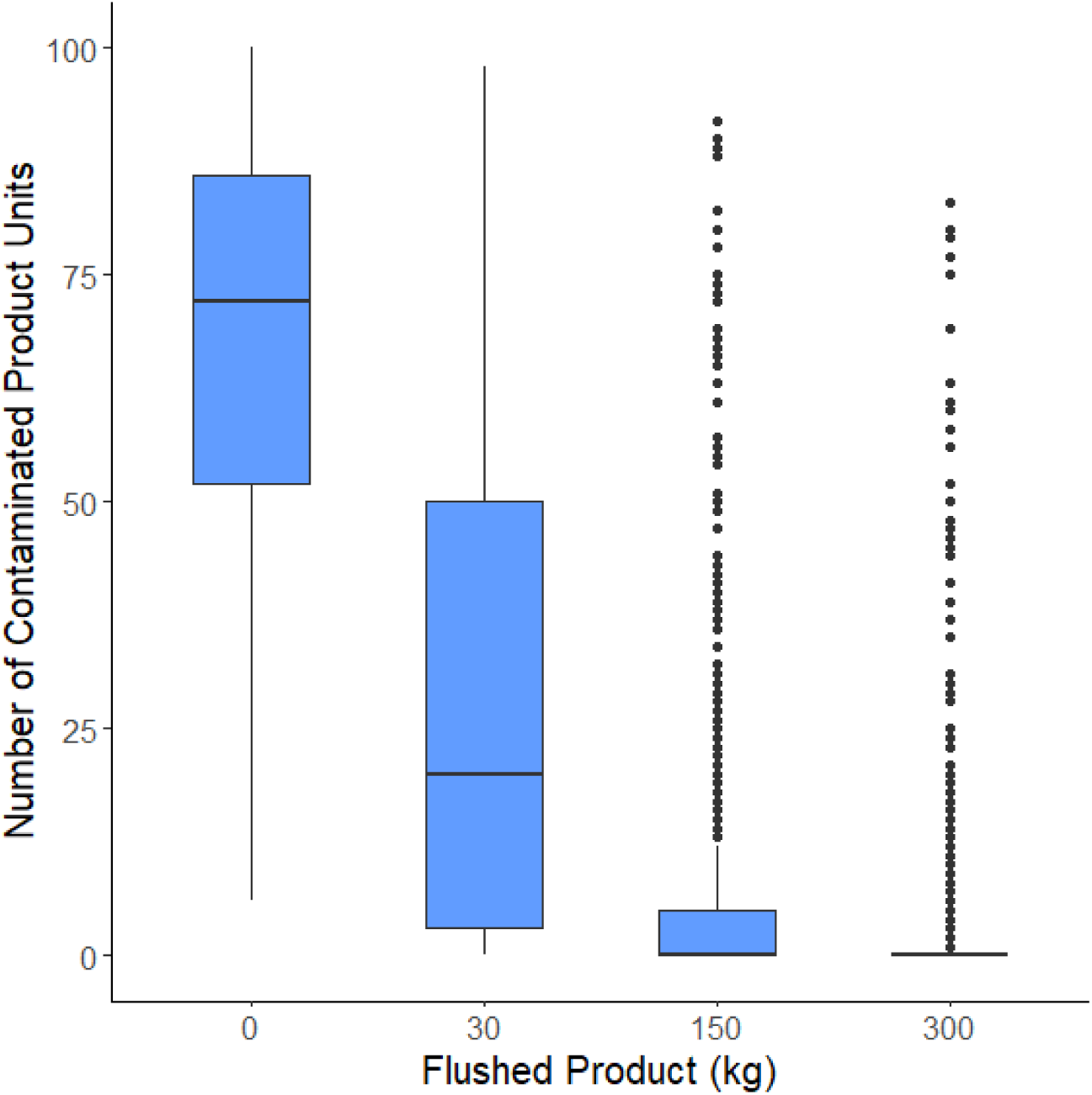
The results of 1,000 simulated production runs for scenario 3 (product flushing) with increasing amounts of product flushed before production begins (30, 150 and 300 kg). *Increasing the amount of flushed product resulted in less product units contaminated with Salmonella. As more product was flushed there was diminishing returns in the reduction of the number of the contaminated product units*.

Increasing the amount of material used in flushing above a certain point provided diminishing returns in the reduction of contaminated units (Fig. 5). Therefore, food safety outcomes must be carefully balanced against increased production costs and food waste. With these considerations, product flushing may be most effective as part of a suite of sanitation strategies and hygienic practices that combine multiple mechanisms of cleaning. Though this approach merits further study to optimize efficacy across key implementation variables including equipment hygienic design, flushing process parameters, flushing material, and surface attributes. As an example, one study found that the rate of transfer from polypropylene surfaces to food product was ineffective for cleaning [37].

Prior research has also indicated the efficacy of flushing. The logarithmic decline in contamination across sequential product units after a breach observed in this study has been previously observed in a bagged lettuce model as well [21]. In this model, transmission of *E. coli* O157:H7 into bagged lettuce was greatest during the initial batches after the breach. Similarly, Muckey et al. found that after introducing a *Salmonella* breach to animal feed mixers, flushing the mixers with two batches of feed resulted in *Salmonella* being below the limit of detection (10 CFU) in both feed and equipment surfaces [27]. In a dry processing environment there may be scenarios when the risk of pathogen growth is elevated such as the presence of residual water after a wet cleaning [46]. Product flushing may be especially effective in these scenarios where there is elevated risk of microbial growth; however, product flushing and other dry cleaning practices are currently not considered a sanitation clean break [12].

### 3.4 Dry wiping with a towel modestly reduced the prevalence and concentration of *Salmonella* contaminated units, with minimal cross-contamination to other surfaces

We simulated dry cleaning the contaminated surface (2 log CFU) by wiping with a dry towel (scenario 3; Fig. 6). This resulted in a 64% reduction in the amount of milk powder units contaminated with *Salmonella* across all simulations: 72 [24, 96] to 26 [12, 64]. The average concentration of *Salmonella* within all contaminated units also decreased from -2.33 log CFU/g [-2.46, -1.86] to -2.42 log CFU/g [-2.48, -2.08]. In a 6 log CFU contamination breach, the percent reduction in the median number of contaminated units from a dry wiping intervention was less (11%): 688 [94, 4420] to 608 [85, 3981], but the reduction in the absolute number of contaminated units (2 log CFU breach: 46 less contaminated units, 6 log CFU breach: 80 less contaminated units) was greater. The average concentration of *Salmonella* within all contaminated units decreased from 0.69 log CFU/g [-0.12, 1.5] to 0.26 CFU/g [-0.65, 1.8]. These results show how the experimentally measured log reductions from towel wiping (< 1 log CFU; Table 1) manifested as modest reductions in the number of *Salmonella* contaminated product units. The reductions in prevalence and concentration of *Salmonella* were less effective compared to flushing for all tested contamination levels (Fig. 6). However, after a 2 log CFU contamination breach, dry wiping still resulted in a 64% reduction in the number of contaminated units, which could be meaningful in reducing downstream human exposure to *Salmonella*. Mechanical dry cleaning methods such as wiping, brushing, and scraping generally offer limited removal of microbial contamination, but can produce visibly clean surfaces [47] that pass ATP tests [48]. As such they are not currently recognized as a validated sanitation method [12], though they are still important sanitation activities for maintaining a hygienic dry processing environment. Our results suggest that rigorous dry physical cleaning has the potential to serve as an effective line of defense against cross-contamination and should be further evaluated.

**Figure 6:**
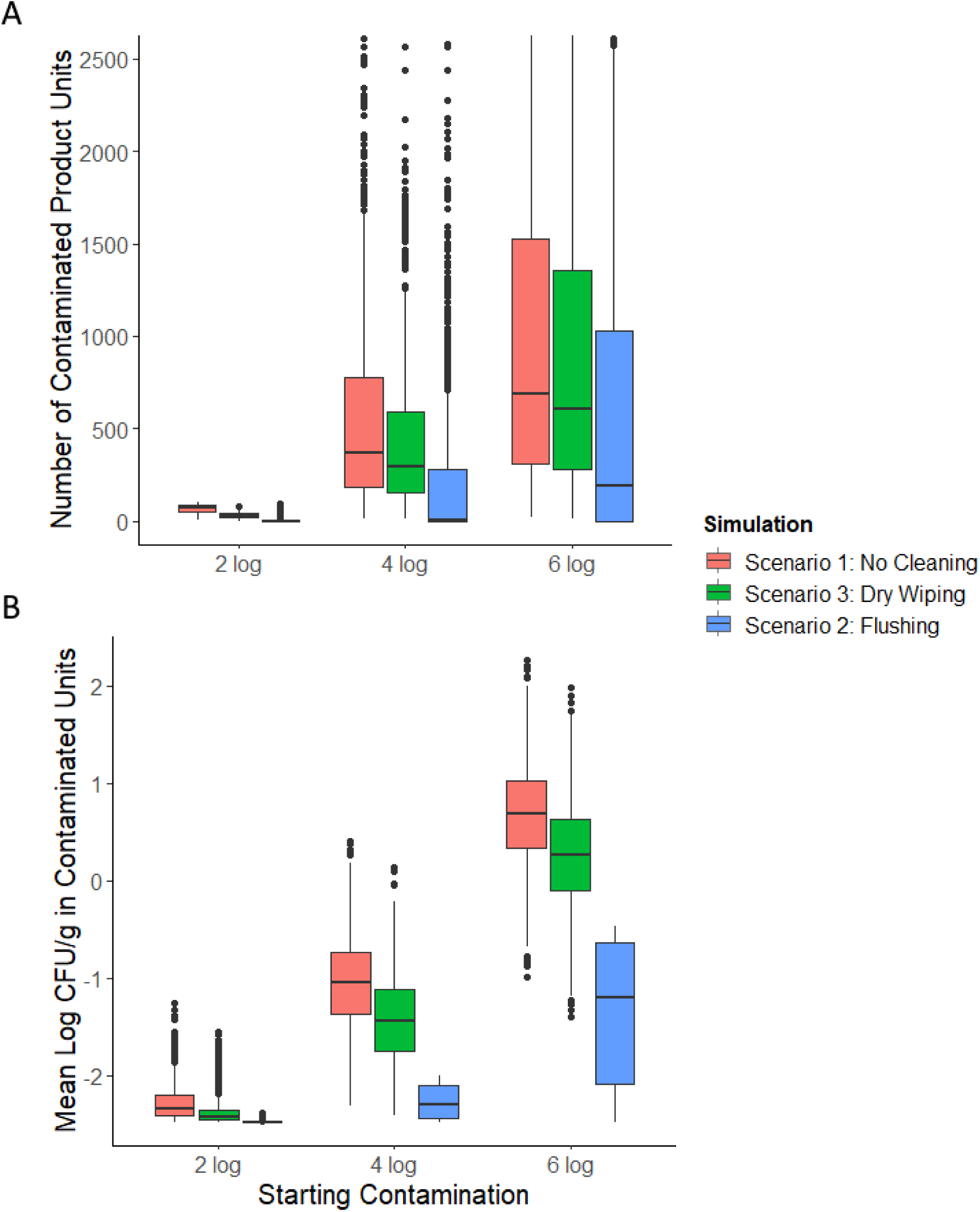
The results of 1,000 simulated production runs for scenario 1, 2 and 3 runs are plotted for each starting contamination level (2, 4, 6 log CFU). **(A)** Each data point represents the number of milk powder units (300 g) contaminated with *Salmonella* in a simulated production run. **(B)** Each data point represents the average concentration of *Salmonella* contamination within the contaminated milk powder units of a simulated production run.

In scenario 4 we modeled a contamination breach where a dry towel becomes contaminated in the process of cleaning a contaminated surface. That towel is then used on a previously clean surface and the *Salmonella* on the towel is transferred to the surface of the processing line (Fig. 1). For this scenario, the towel was previously used on a heavily contaminated surface (6 log CFU). Despite this, the majority of the simulated production runs in scenario 4 (83%) did not result in any cross-contamination to the processing line from the towel. Therefore, downstream contamination to milk powder product was very low. The number of CFU that transferred from the original contaminated surface to the towel and then to the processing line surface was 0 [0, 2]. Out of all simulations, 28 CFU was the largest number of cells transferred from the dry towel to the processing line. These results reflect the low transfer rates from a contaminated surface to a dry towel, and then from a contaminated towel to a clean surface obtained from the experimental data (Table 1).

Cleaning tools are often cited as potential reservoir for contamination in dry processing environments [8], and foodborne pathogens have previously been detected in cleaning tools [49, 50]. Consequently, measures to limit cross-contamination via cleaning tools is considered an important element of GMPs in dry processing facilities [51] [52]. Whether or not cross-contamination of pathogens from cleaning tools is a significant contributor to outbreaks is less clear. A panel of food safety experts rated cleaning tools as medium importance for inclusion in an environmental monitoring program for *Listeria monocytogenes*, and a plurality of experts did not consider cleaning tools to be a transfer reservoir [53]. Our results support the stance that cleaning tools may not be large drivers of cross-contamination, particularly if they are discarded or sanitized between uses, and that the relative risk of other sources in the processing facility may be of more importance to public health. Nonetheless, management of cleaning tools to reduce cross-contamination should still be performed as part of dry processing GMPs [10].

Moreover, research has shown that towels can be a vector for transmission of allergens [54], underscoring the importance of hygienic management.

### 3.5 The surrogate with the most similar cross-contamination dynamics to *Salmonella* varied depending on the scenario

Experimental transfer data was generated for the three potential *Salmonella* surrogates: *E. coli* ATCC 25922, *E. faecium* NRRL B-2354 and *L. innocua* ATCC 51742 (Supplementary file 1) and used to re-run the simulations described in scenarios 1, 2, and 5 (Table 1). Scenario 5 simulated the repeated dry wiping of a contaminated surface until all surface contamination (4 log CFU) was removed and assumed no contamination was re-introduced (Fig. 7 A). Across all simulations, the median number of dry towel cleanings (3 passes of the wipe per cleaning) required to remove all contamination for *Salmonella* was 9 [5, 35]. The most similar surrogate was *L. innocua* which took a median of 12 [6, 64] towel cleanings to remove all the surface contamination. *E. faecium* took only a median of 4 [3, 9] dry cleanings to remove all contamination, which means using it as a surrogate in dry cleaning studies may result in an overestimation of the degree of *Salmonella* reduction. On the other hand, *E. coli* took a median of 20 [10, 101] towel cleanings to remove all contamination. Therefore, *E. coli* would be a very conservative surrogate for removal of *Salmonella* through towel cleaning. The variability in the number of dry wipe cleanings to completely remove all surface contamination should also be considered when choosing a surrogate. While *L. innocua* was the most similar to *Salmonella* in terms of the median number of cleanings to remove all contamination, the variability between simulations was higher (Standard deviation: 20 for *L. innocua* and Standard deviation: 10 for *Salmonella*). This difference in variability introduces uncertainty in the pathogen-surrogate relationship that should be accounted for when testing cleaning interventions.

**Figure 7:**
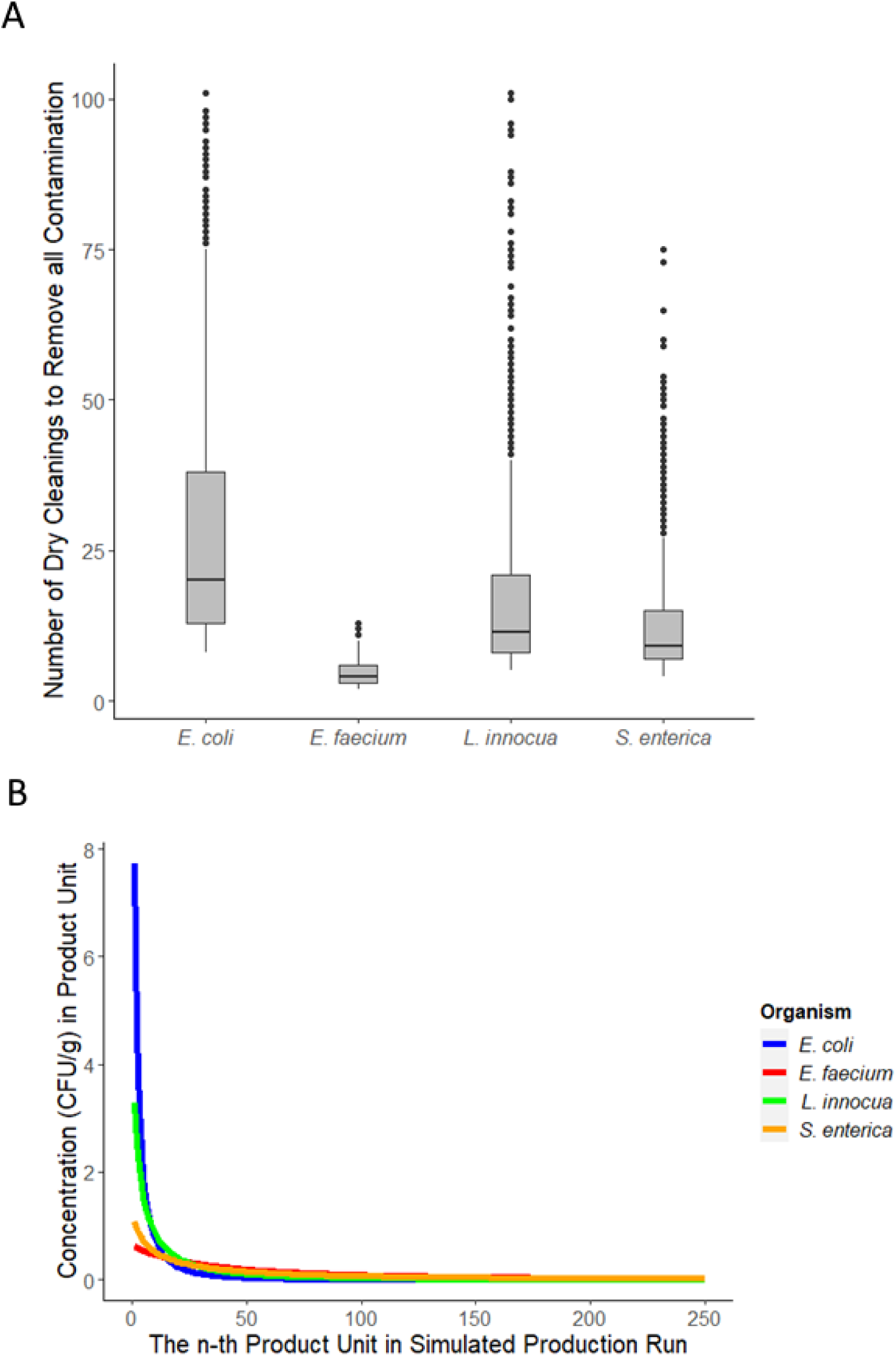
(A) Each data point (n = 1,000 iterations) represents the number of successive cleanings with a dry towel required to remove all surface contamination (4 log) as modeled in scenario 4. *The number of cleanings required was most similar between L. innocua and S. enterica.* **(B)** Across 1,000 simulated production runs (scenario 1, 2 log CFU), the median contamination concentration (CFU/g) in the n-th sequential milk powder unit produced during a production run is graphed for each surrogate. *S. enterica and E. faecium had the most similar diffusion of contamination into product, with less contamination being transferred into each unit but more units becoming contaminated*.

The microbial log reduction (*M_r_*) from dry wiping is the only input parameter in scenario 4, therefore these results reflect the difference in measured log reductions of each bacterium on stainless-steel coupons using a dry towel. For example, *E. coli* had the lowest average log reduction (Table 1) which corresponds to the greatest number of dry towel wipes required to remove all surface contamination. By contrast, in scenario 1 (no dry cleaning intervention), only the transfer coefficient from the contaminated surface into milk powder (*T_c_*) is used as a parameter. In this case, *E. faecium* was the surrogate that behaved most like *Salmonella* (Fig. 7 B). Both showed a pattern of slow transfer of contamination from the surface into milk powder, resulting in more contaminated units with less contamination on average than *L. innocua* and *E. coli*. The number of contaminated units across all scenario 1 simulations for a 2 log CFU contamination breach was 72 [24, 96] for *Salmonella*, 19 [5, 56] for *E. coli*, 74 [44, 93] for *E. faecium*, and 39 [11, 80] for *L. innocua* (Fig. 8 A). By this metric, *E. faecium* resulted in a similar number of cross-contaminated milk powder products as *Salmonella* and a similar variability between simulated production runs. This is in line with the measured transfer rates from the stainless-steel surface to milk powder, which were higher for *E. coli* and *L. innocua* (Table 1). Similarly, the average concentration of *Salmonella* in contaminated units was -2.33 log CFU/g [-2.46, -1.86] which was most similar to *E. faecium* -2.35 log CFU/g [-2.45, -2.12].

**Figure 8:**
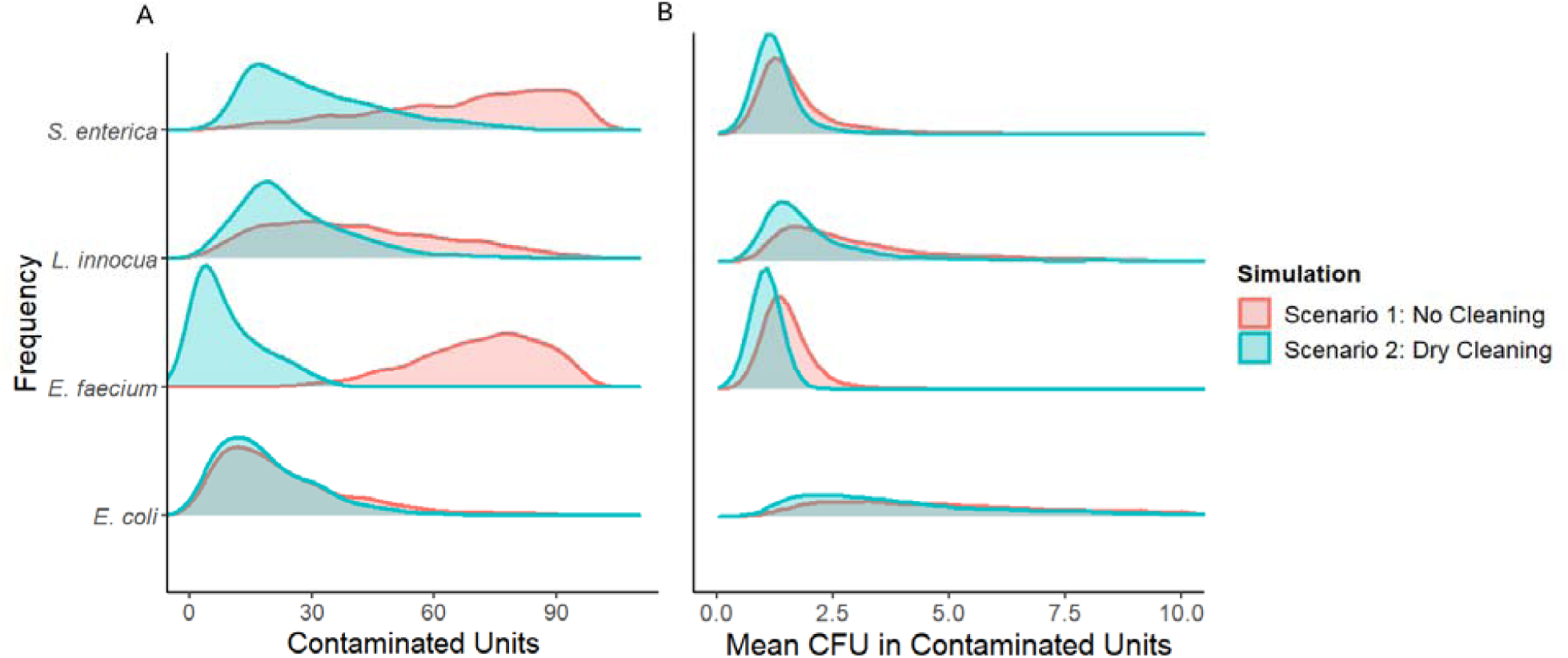
Density estimate of the prevalence of contaminated milk power units **(A)** and the mean concentration within contaminated units **(B)** across all simulated production runs (n = 1,000 iterations) for each organism after a 2 log CFU contamination breach. Higher peaks mean more simulations fell into the range measured on the x-axis. *In terms of contaminated units, Salmonella and E. faecium were most similar in the absence of a towel wiping intervention, but L. innocua was more similar to Salmonella when towel wiping was included*.

In scenario 2 simulations (towel cleaning intervention followed by production), both the dry towel microbial reduction (*M_r_*) and surface to milk powder transfer (*T_c_*) are used as model parameters. In terms of the prevalence of contaminated units, *L. innocua* was the most similar to *Salmonella* (Fig. 8 A). *Salmonella* resulted in 27 [12, 64] contaminated units while *L. innocua* resulted in 22 [8, 53]. *E. faecium* and *E. coli* resulted in 7 [1, 26] and 16 [4, 42] contaminated units respectively in scenario 2. The average contamination concentration in contaminated units was -2.43 log CFU/g [-2.48, -2.08] for *Salmonella* in scenario 2. For *E. faecium*, *L. innocua* and *E. coli* these values were -2.48 [-2.48, -2.38], -2.25 [-2.45, -1.72] and -1.89 [-2.32, -1.28] log CFU/g respectively.

Microbial surrogates have traditionally been used to estimate the lethality of various process controls on relevant foodborne pathogens. In this framework, a good surrogate is characterized by having similar, or slightly more conservative, resistance characteristics to the mode of inactivation in the pathogen of interest under a given control [55]. Additionally, the surrogate should have similar variability in response to the mode of inactivation [55]. *Salmonella* surrogates, such as *E. faecium* NRRL B-2354 [56], have been proposed based on their resistance to thermal inactivation. However, for dry cleaning, the relevant characteristic is transfer and removal during wiping, brushing, scraping, or flushing of surfaces. We found that the surrogate most representative of *Salmonella* depended on the scenario being modeled, and changed as multiple model parameters were considered. For example, in scenario 4 where only dry wiping is considered, the amount of dry towel wipes to remove all contamination from a surface was most similar between *Salmonella* and *L. innocua* (Fig. 7 A), though differences in variability could complicate the pathogen-surrogate relationship. In terms of the number of contaminated milk powder products and the concentration of contamination within those products (scenario 1), *E. faecium* was the most similar to *Salmonella* and had similar variability (Fig. 8 A and 8 B). When a dry wiping intervention was applied before production (scenario 3), *L. innocua* resulted in the most similar number of contaminated product units and similar variability between simulations. Therefore *L. innocua* performed better as a surrogate when the modeled scenario involved a towel wiping intervention, but *E. faecium* performed better as a surrogate when just transfer to milk powder was considered.

Using model outputs as a form of surrogate evaluation can help inform the selection of surrogates used in dry cleaning experiments, modeling applications, and model validation. Direct comparison of empirical transfer and reduction data is often challenging [55]. By contrast, model outputs are more interpretable to industry or regulatory stakeholders and are regularly used as decision support tools. Comparing the predicted number of contaminated units and the concentration of contamination within those units for different surrogates has a clearer connection to the outcome of interest than comparison among, for example, the means of transfer coefficients. An example of this application would be in QMRA style studies that could consider how using transfer data from different *Salmonella* surrogates would affect downstream modeling outcomes like the number of contaminated servings and human exposure. Similarly, how the variability of microbial reduction during dry wiping manifests as a different range in the amount of cleaning required to remove all contamination from a surface gives better context for deciding on microbial surrogates for dry cleaning validation experiments.

As the use of simulation models to assess risk under different sanitation regimes becomes more prevalent, the need for in-plant validation data increases. This is a crucial role for the use of surrogates which may be introduced into commercial and pilot-scale systems. Under some circumstances, for example treatments using milk powder flushing, *E. faecium* may be a suitable candidate in place of *Salmonella*. On the other hand, a validation study for *Salmonella* removal from surfaces through dry wiping should avoid *E. faecium* as a surrogate because it may overestimate the efficacy of the intervention. Rates of cross-contamination are typically influential model parameters [57] and therefore accurate surrogate selection for this parameter is paramount. Surrogate data for transfer rates should be accurate to the medium of interest. In this case we modeled transfer into milk powder, but for other matrices like peanut butter or bulk almonds [58] transfer may be very different.

## CONCLUSION

Overall, these results show the utility of modeling for assessment of dry cleaning strategies and surrogate selection. We demonstrated how empirical values for dry transfer of *Salmonella* into milk powder manifested as prevalence and concentration of contaminated product units in a simulated contamination breach of a processing line. Expanding on this, we showed how dry wiping and product flushing reduced the amount of predicted *Salmonella* contamination. Product flushing, and to a lesser degree dry towel wiping, were effective interventions at reducing the prevalence and concentration of *Salmonella* and should be considered for use in industry in response to suspected contamination breaches or as routine cleaning and sanitation processes. We also demonstrated the value of modeling outputs as a more interpretable basis for microbial transfer surrogate selection. Using this method, we found that the surrogate most appropriate for *Salmonella* dry transfer was dependent on the transfer scenario of interest, underscoring that there is not a one size fits all surrogate for *Salmonella* transfer. Finally, we demonstrated the sensitivity of modeling outputs to important input parameters like surface to product transfer coefficients and the proportion of product that contacts the contaminated surface. Future research should focus on identifying common problem niches and dead zones on processing lines, and measuring the unique cross-contamination dynamics (transfer coefficient during contact, frequency of contact) of these areas so that transfer models may more accurately represent real world conditions. Future modeling applications may also consider the trade-off in enhanced sanitation and microbial growth potential that occurs during wet cleaning and how these risks compare to exclusive dry cleaning.

## Data Availability

All data produced in the present work are contained in the manuscript

